# The Exodus of Healthcare Professionals from Ghana and its Effect on the Healthcare System

**DOI:** 10.1101/2025.05.10.25327355

**Authors:** Julius Caesar Mahama, Daphne Kleopa, Prince Yeboah Kumi, Simon Nyarko

**Affiliations:** School of Health Sciences, Liverpool John Moore University, UK; School of Medicine, University for Development Studies, Tamale, Ghana; Department of Pharmaceutics, Kwame Nkrumah University of Science and Technology, Kumasi, Ghana

**Keywords:** Brain drain, Healthcare workforce migration, Health workforce shortages, Ghana, Scoping review

## Abstract

The migration of healthcare professionals from low- and middle-income countries (LMICs) to high-income countries (HICs), often termed “brain drain,” remains a significant and widely debated issue. This study examines the exodus of healthcare workers from Ghana and its impact on the country’s health system. Using a scoping review methodology, we mapped and synthesized existing literature on healthcare professionals’ migration from Ghana, focusing on key challenges and strategies to address this phenomenon. The Population, Concept, Context (PCC) framework was systematically applied to define study eligibility criteria. The search strategy employed Boolean operators (AND/OR) with key MeSH words including healthcare workforce, health policy, migration, brain drain, intervention strategies, and Ghana across databases such as PubMed, Google Scholar, Scopus, and Web of Science. Eleven (11) studies met the inclusion criteria. The review identified low wages, poor working conditions, limited career advancement opportunities, and political instability as primary drivers of healthcare professionals’ migration. To mitigate these challenges, efforts should focus on improving domestic conditions for healthcare workers, including enhanced remuneration, better service conditions, and career development initiatives. Additionally, governments must prioritize strengthening healthcare infrastructure, particularly in rural regions, to reduce disparities in service delivery. The findings also suggest that partnerships with high-income countries could facilitate skills development programs for Ghanaian healthcare workers, provided such collaborations include strengthened agreements (e.g., bonding schemes) to ensure professionals return to apply their expertise domestically. While the scoping review synthesizes critical insights, further empirical studies targeting specific regions or demographic blocs are needed to assess whether migration trends have persisted or evolved. Future research should also explore new subpopulations, geographic contexts, or timeframes to evaluate the reproducibility of existing findings and their applicability to contemporary or shifting socioeconomic landscapes.

## Introduction

The trend of healthcare professionals migrating from low- and middle-income countries (LMICs) to high-income countries (HICs), often referred to as the “brain drain”, has been a longstanding point of discussion in recent times. Migration is a complex process and can be either a necessity or choice driven by various factors, both negative (push factors) and positive (pull factors), leading individuals to seek better opportunities and living conditions. The motivations behind this decision can be economic, political, cultural, professional, personal, or a mix of these factors [1].

The exodus of healthcare professionals from Ghana, often referred to as “brain drain,” has significant implications for the country’s healthcare system. This phenomenon is driven by multiple factors, including inadequate remuneration, poor working conditions, and limited professional development opportunities, which collectively contribute to a high turnover intention among healthcare workers [2]. The migration of healthcare professionals has been documented to result in a scarcity of skilled personnel, weakened healthcare systems, and substantial economic losses, with estimates suggesting that Ghana loses approximately $415 million annually due to physician migration [3].

The healthcare system in Ghana has seen significant improvement over the years. This positive transformation is largely due to the annual increase in the number of healthcare professionals, coupled with the provision of essential medical equipment in health institutions [4]. Every year about 600 medical officers and nurses graduate from Ghana’s nursing and medical schools and are subsequently absorbed into the country’s healthcare system [5]. Despite these encouraging prospects for the healthcare system, the ratio of patients to healthcare professionals in Ghana remains below the WHO recommended [6].

Research indicates that dissatisfaction among healthcare workers is closely linked to their working conditions. A study highlighted that low salaries, inadequate opportunities for continuing professional education (CPE), and poor equipment significantly influence the decision of healthcare professionals to seek employment abroad [7]. Furthermore, the lack of structured support systems and professional development pathways exacerbates the situation, leading to a workforce that is not only demotivated but also underprepared to meet the healthcare needs of the population [8]. The absence of gerontology specialists, for instance, reflects a broader issue of inadequate training and specialization within the healthcare workforce, which is critical as Ghana’s population ages [9], [10].

Moreover, the implications of this migration extend beyond immediate healthcare delivery challenges. The loss of healthcare professionals undermines the quality of care available to vulnerable populations, particularly in rural areas where access to healthcare is already limited [11]. The migration of skilled workers also leads to increased reliance on less experienced personnel, which can compromise patient safety and the overall effectiveness of healthcare services [12]. Consequently, the healthcare system in Ghana faces a dual challenge: addressing the immediate impacts of workforce shortages while simultaneously implementing strategies to retain existing staff and attract new talent.

The effects of healthcare professional migration are essential for the Ghanaian government and healthcare authorities to prioritize improvements in working conditions, including competitive remuneration, opportunities for professional development, and supportive workplace environments. Initiatives aimed at enhancing job satisfaction and motivation among healthcare workers are crucial for retention [13]. Additionally, policies that promote equitable distribution of healthcare professionals, particularly in underserved areas, can help address the disparities in healthcare access and quality [14].

According to a statistics from the Ministry of Health (MOH), roughly 34% of medical professionals—including physicians and nurses—who received training between 2008 and 2015 departed Ghana to find employment abroad [15]. According to these figures, Ghana’s medical workforce is seriously underperforming and needs immediate policy attention. The Ghana Health Service has revealed that 3,688 medical professionals have left the nation in search of better opportunities overseas over the last three years, both crucial and expert careers. Some healthcare professionals left Ghana because of poor working conditions, which prompted them to look for better opportunities abroad. The healthcare systems of low-income nations, especially those in sub-Saharan Africa, including Ghana, are seriously threatened by the increased international migration of health professionals [16].

A significant threat to Ghana’s efforts to achieve health equity is the exodus of medical professionals, particularly doctors and nurses [17]. Significant research gaps persist in our understanding of the migration of skilled healthcare professionals from Ghana. Comprehensive and reliable information remains elusive, particularly in areas such as the push and pull factors influencing this migration, the impact of this exodus on the healthcare system, the primary countries these professionals are migrating to, and the strategies implemented by the Ghanaian government to mitigate this escalating brain drain. These areas are critical for a thorough understanding of the situation and for the development of effective interventions.

The exodus healthcare workers from Ghana have a dire consequence on the Ghanaian health sector. It is very unfortunate and concerning to know that, developing nations, with their limited resources, would invest in the medical and nursing education of their citizens with the hope of improving their healthcare systems, only to lose them to the West and Europe. As a result, the aim of the study is to examine the existing literature on the exodus of healthcare professionals from Ghana and its impact on the healthcare system in the country. It therefore sought to find answers to these questions; What is the extent of the exodus of healthcare professionals from Ghana? what are the causes of the exodus of healthcare professionals from Ghana? what are some of the effects of the exodus of healthcare professionals on the healthcare system in Ghana? and in what ways can the exodus of healthcare professionals from Ghana be reduced?

The current body of literature on this subject is relatively sparse and has not been systematically searched, summarized, or synthesized. Therefore, a scoping review is essential to understand the totality of this literature, including the interventions and guidelines that have been implemented, the causes and impact/effects on the continuous exodus of healthcare workers on the healthcare system in Ghana, and the gaps in knowledge that still exist. This review will provide a comprehensive understanding of the situation and inform future strategies to address this critical issue.

## Methods

### Study Design

This study employed a scoping literature review methodology, analyzing academic literature that had undergone rigorous peer review and was published in respected scientific journals. It incorporated various study designs, including randomized controlled trials (RCTs) and observational studies (cohort, case-control). The scoping review was conducted by systematically searching reference lists of previously published studies and reviews to identify relevant papers. This approach allowed for a holistic synthesis of available studies, identifying key trends, causes, consequences, and possible interventions.

### Setting

The research focused on Ghana, a lower-middle-income country facing significant healthcare workforce challenges due to migration. Studies included in the review examined healthcare professionals’ migration patterns, motivations, and systemic impacts within the Ghanaian healthcare landscape.

### Participants

The study did not involve direct human participants, as it was based on secondary data extracted from published academic literature, governmental reports, and healthcare workforce databases. The review covered several key databases, including PubMed, Scopus, Google Scholar, Embase, CINAHL, PsycINFO, and Web of Science, to ensure a comprehensive inclusion of significant studies, minimizing the risk of missing relevant research.

### Variables

The key variables analyzed included:

- Migration trends of healthcare professionals from Ghana
- Economic, professional, and systemic drivers of migration
- Effects of migration on healthcare service delivery
- Policy interventions addressing workforce retention

### Data Sources/Measurement

A systematic search was conducted across databases including PubMed, Google Scholar, Scopus, and Web of Science. Boolean operators (AND/OR) were utilized alongside MeSH terms such as “healthcare workforce,” “brain drain,” “migration,” “intervention strategies,” and “Ghana.” Relevant studies were screened using title, abstract, and full-text review, following a predefined eligibility framework.

### Bias

The PCC framework (Population, Concept, Context) guided the development of the eligibility criteria, focusing on the extent, causes, and effects of healthcare professionals’ exodus, as well as potential solutions. Comparison groups were selected based on emerging issues related to the exodus. To minimize selection bias, studies were included only if they met rigorous inclusion criteria based on relevance, methodological quality, and applicability to the Ghanaian healthcare context. Conference abstracts, unpublished reports, and non-peer-reviewed literature were excluded to ensure methodological consistency. The quality of the studies was assessed using the Newcastle-Ottawa Scale (NOS) for observational studies and the Cochrane Risk of Bias Tool for RCTs. These tools evaluated key areas such as randomization, blinding, allocation concealment, and reporting biases. The risk of bias was classified as low, moderate, or high based on these assessments, following the Cochrane Handbook for Reviews of Interventions.

### Study Size

A total of 11 studies met the inclusion criteria, providing a comprehensive representation of healthcare migration trends affecting Ghana.

### Quantitative Variables

Where applicable, the study analyzed numerical indicators such as healthcare workforce migration rates, financial losses due to migration, and shortages in specific healthcare roles.

### Statistical Methods

Data extracted from the studies were synthesized using descriptive statistical summaries and thematic mapping. Quantitative results were reported through frequency distributions, percentages, and comparative trend analyses.

## Results

Table 1, 2 and 3 summarized and synthesized the findings from existing studies on the subject matter. Table 1 highlights the studies on health workers and the effects of migration, outcomes and job satisfaction. Table 2 highlights the methodological approaches used in interventional studies, and Table 3 highlights the interventions outlined in these studies

**Table 1:** Studies on health workers and the effect of health and job interventions on outcomes and improvement of remuneration.

| Author, Year | Research Context |  |  |  |  |
| --- | --- | --- | --- | --- | --- |
|  | Assessment Type | Recommended Intervention | Population of Interest | Outcome | Job Satisfaction |
| (Adjei-Mensah, 2023) | Survey | Increase remuneration and motivation of workers | Health workers | Brain drain | Low remuneration |
| (Wutor et al., 2024) | Survey | Improve remuneration & enhance career development | Medical doctors | - | Low remuneration |
| (Sakeah et al., 2023) | Quasi-experimental study | Prioritising domestic healthcare systems and offering competitive incentives | Health workers | Migration | Low remuneration |
| (Ohene-Botwe et al., 2024) | Survey | Financial incentives, career development and improved conditions | Radiographers | - | Low remuneration |
| (Asamani et al., 2021) | Case study | Redistribution of the health workers | Health workers | Lack of human resources/ health workers | - |
| (Poku et al., 2023) | Cross sectional study | Development of assistance and promoting good governance | Health workers | Migration | Low remuneration |
| (Boadu et al., 2021) | Report | Provision of better working conditions, competitive remuneration and motivation, and opportunities for professional development | Health workers | - | Low remuneration |
| (Oduola, 2023) | Systematic review article | Viable health and policy approaches | Health workers | Brain drain | Low remuneration |
| (Adua et al., 2017) | Review article | Research report | Health workers | Lack of health infrastructure | - |
| (Ibrahim et al., 2024) | Qualitative study | Career advancement opportunities and improving salaries of health workers | Health workers | - | Low remuneration |
| (Akinto, 2021) | Review article | Increase the salary of health workers | Health workers | - | Low remuneration |

**Table 2:** Methodological Approaches used in interventional studies for job satisfaction.

| **Paper Source** | **Study design** | **Methodology type (tool used)** | **Sample Size** |
| --- | --- | --- | --- |
| (Adjei-Mensah, 2023) | Empirical study | Quantitative /Survey | 400 |
| (Wutor et al., 2024) | Cross sectional study | Quantitative /Survey | 641 |
| (Ibrahim et al., 2024) | Systematic review | Qualitative /Secondary data | NA |
| (Ohene-Botwe et al., 2024) | Cross sectional study | Quantitative / Survey | 128 |
| (Asamani et al., 2021) | Systematic review | Qualitative / Secondary data | NA |
| (Poku et al., 2023) | Cross sectional study | Quantitative / Survey | 225 |
| (Amoo et al., 2024) | Systematic review | Qualitative / Secondary data | 6 news agencies on nurses |
| (Oduola, 2023) | Systematic review | Qualitative / Secondary data | NA |
| (Adua et al., 2017) | Systematic review | Quantitative / Secondary data | NA |
| (Ibrahim et al., 2024) | Empirical study | Qualitative / Interview guide | 16 |
| (Akinto, 2021) | Systematic review | Qualitative / Secondary data | NA |

**Table 3:** Interventions outlined in the reviewed papers.

| **Studies** | **Intervention** | **Benefits of intervention** |
| --- | --- | --- |
| (Adjei-Mensah, 2023; Boadu et al., 2021; Wutor et al., 2024) | Increased remuneration and motivation of staff | High remuneration is beneficial in reducing brain drain |
| (Ibrahim et al., 2024; Ohene-Botwe et al., 2024) | Enhanced career development/advancement of career | Not entirely beneficial because career development does not necessarily lead to high remuneration or improve condition of service |
| (Adua et al., 2017; Akinto, 2021; Boadu et al., 2021; Ibrahim et al., 2024) | Prioritising domestic healthcare, offering competitive incentives and increase in salary | High remuneration is beneficial in reducing brain drain |
| (Oduola, 2023) | Viable health and public policy approaches | Effective health/public policy can improve condition of service |
| (Asamani et al., 2021) | Redistribution of the health workers | Beneficial in narrowing the staffing gap |
| (Poku et al., 2023) | Promoting good governance | Beneficial in improving health care outcome |

### 3.1 Migration Trends

The migration of health professionals from Ghana to other countries, often referred to as “brain drain,” has been significant over the past few decades, with various studies highlighting the extent and consequences of this phenomenon (see Table 1). Empirical literature shows that Ghana, like many sub-Saharan African countries, has faced a persistent exodus of healthcare workers, particularly doctors and nurses, to countries with better working conditions, higher salaries, and more opportunities for professional development [18]. Ghana lost over 50% of its medical graduates to more developed countries like the United Kingdom, United States, and Canada between 2009 and 2021, which severely strained the country’s health system, leading to inadequate staff levels in many healthcare facilities [19]. Similarly, the World Health Organization (WHO) reports that in 2015, nearly 3,000 Ghanaian-trained physicians and over 2,000 nurses were working abroad, mostly in high-income countries [20]. This represents a significant portion of Ghana’s trained healthcare professionals, thus exacerbating shortages in the domestic healthcare system.

### 3.2 Drivers of Migration

The migration of health professionals from Ghana, commonly referred to as “brain drain,” is driven by a combination of economic, professional, and systemic factors. Empirical literature highlights several key causes (see Table 1). Economic factors, for instance, are among the most frequently cited reasons. Low wages, poor working conditions, and a lack of financial incentives in Ghana’s healthcare system push professionals to seek better-paying opportunities abroad. Studies by Boadu [17] reveal that significant salary differentials between Ghana and countries like the United States, United Kingdom, and Canada are major pull factors, as professionals can earn significantly more abroad. Furthermore, poor working conditions and inadequate resources in Ghana’s healthcare system exacerbate this migration. Health professionals often face challenges such as inadequate medical supplies, poorly equipped facilities, and limited opportunities for career advancement [21]. As a result, doctors, nurses, and pharmacists, frustrated by their inability to provide optimal care, are compelled to seek out countries with better healthcare infrastructure [22]. Professional development and educational opportunities also play a significant role in this migration. Many healthcare professionals in Ghana feel that opportunities for further education, specialization, and career growth are limited [23]. Countries with more advanced healthcare systems offer structured postgraduate training and specialization programs that appeal to professionals eager to advance their careers, particularly doctors and nurses [24].

Additionally, political and systemic factors, such as instability in health sector policies, lack of job security, and slow bureaucratic processes, further fuel migration. Studies suggest that frequent political changes lead to policy inconsistencies, demoralizing healthcare workers and creating dissatisfaction within the system [18]. Moreover, the perceived undervaluation of health professionals, evidenced by inadequate recognition and delayed promotions, accelerates migration. Finally, social and personal factors, such as family reunification, lifestyle choices, and the desire for safety, also play a role. Many health professionals, seeking a higher standard of living and better social services, are influenced by personal aspirations and family ties abroad [25]. In a nutshell, economic factors, including low wages and inadequate working conditions, were identified as major push factors. Poor healthcare infrastructure, limited career advancement, and political instability also contributed to migration trends.

### 3.3 Effects on the Healthcare System

The migration of health professionals from Ghana to other countries, often referred to as the “brain drain,” has significant consequences for the country’s healthcare system, economy, and overall development. One of the most critical effects is the shortage of skilled healthcare workers, particularly in rural and underserved areas. The emigration of doctors, nurses, and other health professionals creates gaps in the delivery of essential healthcare services, leading to increased workloads for the remaining staff and a decline in the quality of care [26]. This shortage further weakens an already fragile healthcare infrastructure, making it difficult to meet the population’s healthcare needs, especially for diseases such as HIV/AIDS, malaria, and tuberculosis. Moreover, the migration of healthcare professionals impacts medical training and education in Ghana. As experienced professionals leave, the capacity of local medical institutions to educate and mentor future health workers diminishes, creating a cycle that hinders the development of healthcare providers and ultimately weakens the healthcare system [23].

In addition to its effect on healthcare delivery, the migration of health professionals has a notable impact on Ghana’s economy. The country invests substantial resources in training healthcare workers, but when these professionals leave for better opportunities abroad, this investment is effectively lost. Asamani and colleagues estimate that educating a single doctor in Ghana can cost over $60,000, with the receiving countries benefiting from the investment without incurring the training costs [26]. This “reverse subsidy” deprives Ghana of its human capital while benefiting wealthier nations. However, some studies suggest potential benefits, such as remittances sent home by health professionals working abroad, which can contribute to the local economy by supporting families and funding education [17]. Additionally, international experience and skills gained abroad may benefit Ghana if these professionals return or contribute through knowledge transfer. Nevertheless, the overall impact remains negative, as the loss of skilled healthcare workers continues to strain the healthcare system and impede development efforts (see Table 3).

### 3.4 Intervention Strategies

Several interventions have been proposed and implemented to manage the migration of health professionals from Ghana to other countries, with empirical literature highlighting strategies at the national, institutional, and international levels (see Table 3). One key intervention is improving working conditions in the health sector, as poor working environments, inadequate remuneration, and limited career development opportunities are major factors driving the migration of health professionals [20]. Addressing these issues through salary increases, improved infrastructure, and continuous professional development programs can reduce the desire to migrate. For instance, Boadu (2021) notes that reforms targeting better pay and workplace conditions in Ghana have been modestly successful in retaining healthcare workers, although challenges remain. Moreover, bonding schemes where newly trained health professionals are required to work in the country for a set period before being eligible to emigrate have been used to curb immediate outflow. However, Ibrahim et al. (2024) suggest that these schemes are only effective when pull factors, such as better opportunities abroad, are also addressed.

International partnerships and agreements also play a vital role in managing migration. The UK and Ghana, for example, have engaged in bilateral agreements that facilitate temporary migration for skills development while ensuring that health professionals return after a specified period [27]. Additionally, encouraging task-shifting within the health sector has been shown to alleviate the impact of migration. Task-shifting allows less specialized healthcare workers to take on roles traditionally performed by highly trained professionals, ensuring continuity of care despite workforce shortages [3].

Finally, retention strategies focusing on incentives like rural allowances, housing benefits, and access to further education are crucial in motivating health professionals to remain in the country, particularly in underserved areas [18]. These interventions, when applied holistically, can significantly reduce the migration of health professionals from Ghana.

## Discussion

The results presented provide a comprehensive understanding of the extent, causes, and consequences of the migration of health professionals from Ghana, as well as interventions aimed at managing this persistent issue. The significant exodus of health professionals, particularly doctors and nurses, has been well-documented, with empirical literature highlighting the severe impact on the healthcare system (see Table 1). The loss of over 50% of Ghana’s medical graduates to developed countries between 2009 and 2021, as Adjei-Mensah in 2023 reveals, illustrates the magnitude of the brain drain. Similarly, another study reported on thousands of Afro-trained physicians and nurses working abroad further underscores how this migration exacerbates shortages in healthcare personnel, especially in rural areas [28].

The results show that the migration is driven by a combination of push and pull factors. Poor remuneration, lack of career advancement opportunities, and inadequate infrastructure in Ghana serve as strong push factors, while higher wages, better working conditions, and professional development opportunities abroad act as pull factors. These findings are consistent with studies by [29] and Darko-Gyeke et al [30], which emphasize the disparity in working conditions between Ghana and receiving countries like the United States and the UK. Moreover, aggressive recruitment policies in developed countries further fuel the exodus, as highlighted by Buchan et al [31], resulting in a substantial depletion of the health workforce.

The impact of this migration is seen not only in the immediate loss of skilled workers but also in the long-term weakening of the healthcare system’s capacity. The reduction in available mentors and educators, as discussed by Toyin-Thomas et al [32], has significant implications for the training of future professionals, creating a cycle that perpetuates the shortage of healthcare workers. Additionally, the economic impact is considerable, with Ghana losing significant investments in the training of health professionals. Sadiq and colleagues [33] highlight the financial burden of educating health professionals who ultimately emigrate, emphasizing the detrimental effects of this “reverse subsidy.”

The interventions suggested such as improving working conditions, implementing bonding schemes, and fostering international partnerships are necessary but not without challenges. While efforts to enhance pay and working conditions have had some success, as [20] notes, retention remains difficult without addressing external pull factors. The mixed effectiveness of bonding schemes, as pointed out by Hussain and peers [34], indicates that these interventions alone are insufficient. Furthermore, task-shifting and retention strategies, particularly in rural areas, show promise in mitigating the impact of migration, as noted by Mundeva et al [35]. However, these strategies must be part of a broader, holistic approach that addresses both the push and pull factors driving health professionals out of Ghana.

## Conclusion

The exodus of health professionals from Ghana presents a critical challenge to the country’s healthcare system, impacting service delivery, healthcare access, and the economy. This brain drain, characterized by the departure of doctors, nurses, and other healthcare workers to more developed countries, has strained Ghana’s health infrastructure, especially in rural and underserved areas. The review found that over 50% of Ghana’s medical graduates emigrated to high-income countries between 2009 and 2021. Healthcare workforce depletion was particularly significant in rural and underserved areas. The primary causes driving this migration include low wages, inadequate working conditions, limited career advancement opportunities, and political instability. High-income countries continue to attract Ghanaian health workers with better pay, advanced facilities, and structured professional development programs. To mitigate this crisis, Ghana requires a multipronged approach focusing on improving domestic conditions for healthcare professionals and establishing frameworks to manage migration sustainably. First, increasing salaries, enhancing workplace environments, and providing consistent professional development opportunities are essential steps to retain healthcare workers. The government could prioritize investments in healthcare infrastructure, especially in rural regions, where the shortages are often most severe. Second, retention strategies like rural allowances, housing benefits, and continuous education incentives can encourage health professionals to remain within the country.

On a policy level, Ghana could benefit from international partnerships that allow for skills development abroad while ensuring that professionals return to contribute locally. Bonding schemes, while partially effective, may be improved through collaborations with high-income countries to offer temporary migration options without losing professionals permanently. Additionally, expanding task-shifting initiatives, wherein less specialized staff are trained to perform essential roles, can help offset the impact of migration by ensuring continuity of care even with fewer personnel.

The migration of skilled healthcare professionals weakened service delivery, particularly specialized care areas such as gerontology and primary healthcare. Increased workloads, reduced mentorship opportunities, and systemic healthcare inefficiencies were among the consequences. Ultimately, addressing Ghana’s brain drain will require a coordinated approach across government, healthcare institutions, and international partners. By improving working conditions, strengthening policy frameworks, and creating development pathways, Ghana can better retain its skilled health workforce, promoting a sustainable and resilient healthcare system. Proposed measures included improved remuneration, better career progression frameworks, and structured agreements between Ghana and high-income countries to manage temporary workforce migration while ensuring skills retention.

The findings of this study however, is based on existing studies. In as much as this methodology is accepted in scientific studies, it is important to engage in empirical studies with focus on a particular region or bloc to assess whether the exodus of healthcare professionals has changed or otherwise. Therefore, further studies on related subject with a new or sub population or audience with different timeframe is required to assess whether existing studies are reproducible to new population and applicable to different era

### Limitations of the study

The quality of the included studies varies significantly, with some exhibiting methodological limitations such as small sample sizes, self-reported data, or lack of longitudinal analysis. These inconsistencies may affect the robustness and reliability of the synthesized findings. Also, the review’s scope is restricted to Ghana, which limits its generalizability to other low- and middle-income countries with distinct socioeconomic, political, or healthcare contexts.

### What is known about this topic

- Low wages, poor working conditions, and limited career advancement opportunities are primary drivers of healthcare professionals’ migration from Ghana.
- The exodus exacerbates workforce shortages, strains healthcare delivery (especially in rural areas), and results in significant economic losses due to “reverse subsidies” from training investments.
- Proposed interventions include improving remuneration, enhancing career development pathways, and fostering international partnerships with bonding schemes.

### What this study adds

- A comprehensive synthesis of existing literature using the PCC framework, mapping the extent, causes, effects, and interventions related to healthcare professionals’ migration from Ghana.
- Identification of critical research gaps, such as the need for empirical studies on regional disparities, evolving migration trends, and the effectiveness of bonding schemes in retaining professionals.
- A holistic policy recommendation framework emphasizing coordinated domestic reforms (e.g., rural incentives, infrastructure upgrades) and equitable international collaborations to address both push and pull factors.

**Figure 1.**
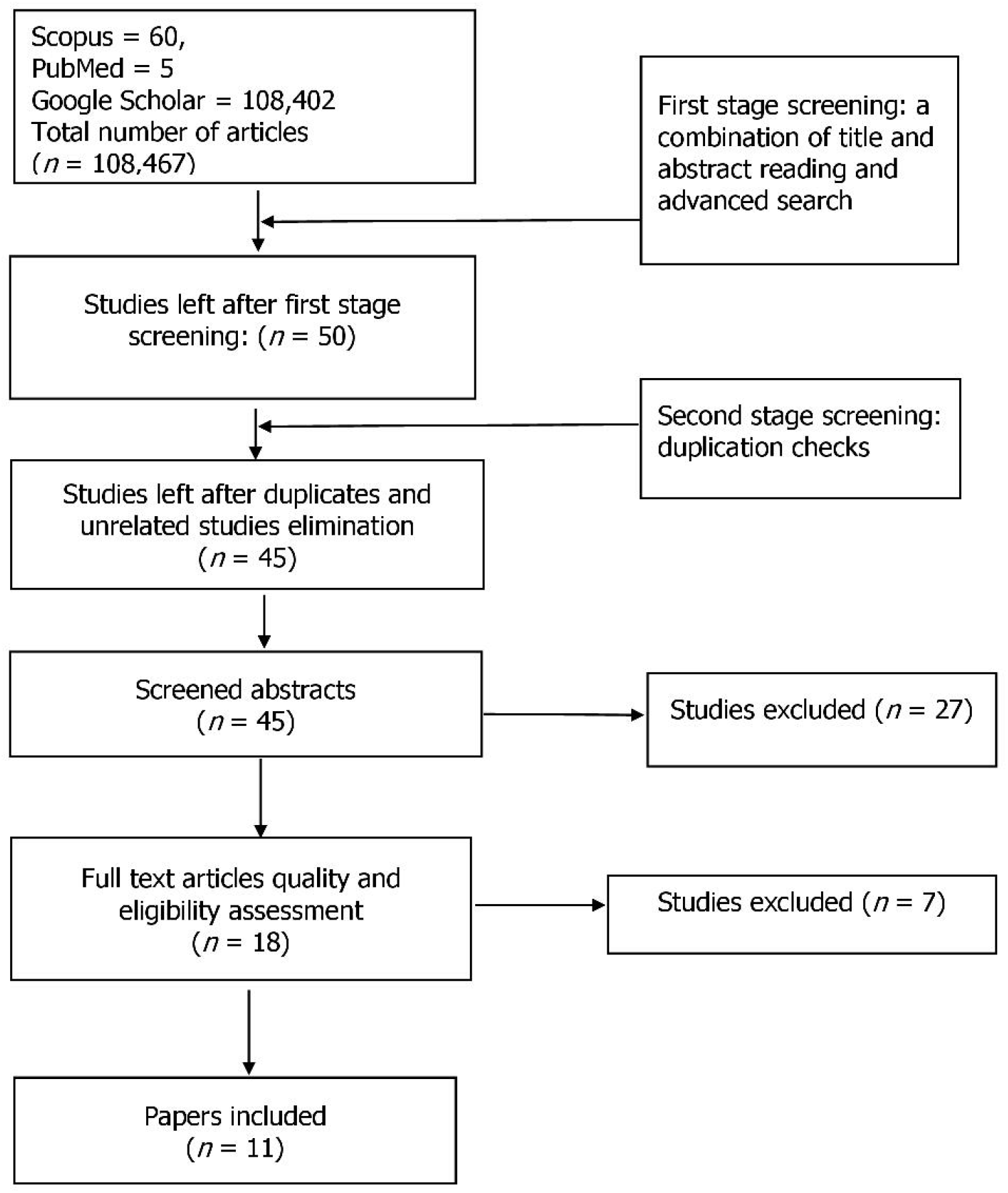

## Data Availability

All data produced in the present work are contained in the manuscript

